# Determining prescriptions in electronic healthcare record (EHR) data: methods for development of standardized, reproducible drug codelists

**DOI:** 10.1101/2023.04.14.23287661

**Authors:** Emily L Graul, Philip W Stone, Georgie M Massen, Sara Hatam, Alexander Adamson, Spiros Denaxas, Nicholas S Peters, Jennifer K Quint

## Abstract

**Objective:** Epidemiological research using electronic healthcare records(EHR) informing everyday patient care uses combinations of codes (“codelists”) to define diseases and prescriptions (or phenotypes). Yet methodology for codelist generation varies, manifesting in misclassification bias, while there are drug-specific codelist considerations.

**Materials and Methods:** We developed methods to generate drug codelists, testing this using the Clinical Practice Research Datalink (CPRD) Aurum database, accounting for missing data in “attribute” search variables. We generated codelists for 1)cardiovascular disease and 2)inhaled Chronic Obstructive Pulmonary Disease (COPD) therapies, applying them to a sample cohort of 335,931 COPD patients. We compared searching on all search variables (A,”gold standard”) to B) chemical and C) ontological information only.

**Results:** In Search A we determined 165,150 patients prescribed cardiovascular drugs(49.2% of cohort), and 317,963 prescribed COPD inhalers (94.7% of cohort). Considering output per value set, Search C missed substantial prescriptions, including vasodilator anti-hypertensives (A and B:19,696 prescriptions; C:1,145) and SAMA inhalers (A and B:35,310; C:564).

**Discussion:** We recommend the full methods (A) for comprehensiveness. There are special considerations when generating adaptable and generalizable drug codelists, including fluctuating status, cohort-specific drug indications, underlying hierarchical ontology, and statistical analyses.

**Conclusions:** Methods must have end-to-end clinical input, and be standardizable, reproducible, and understandable to all researchers across data contexts.

**LAY ABSTRACT:** Health research using patient records informs everyday medicine, using groups of codes (“codelists”) to define diseases and drugs. Yet methods to create drug codelists are inconsistent, may not include physician expertise, nor be reported.

We developed a reproducible method to create drug codelists, testing it using de-identified healthcare records. We generated codelists for 1) heart conditions and 2) inhalers to identify prescriptions in a sample group of 335,931 patients with chronic lung disease. We compared our full methods (Search A) to two restricted searches to show prescriptions can be missed if necessary considerations are not made.

In search A, we determined 165,150 people (49.2% of sample group) prescribed drugs from the heart codelist. For lung inhalers, we determined 317,963 prescriptions (94.7% of group). Search C missed substantial prescriptions, for drugs lowering blood pressure by opening vessels (A and B:19,696 prescriptions; C: 1,145), and short-term inhalers opening airways (A and B: 35,310; C:564).

We recommend full methods(A) for completeness. Drug codelist methods must be consistent, duplicable, and include physician input at all research stages, and have special considerations including status (eg, new, taken off market), disease, and drug categorical system. Quality methods should be freely accessible and usable across study contexts.

## INTRODUCTION

### Health data research and codelist generation

Research using electronic health records (EHR) is increasingly used to inform patient care across a breadth of longitudinal data sources, including the U.S. Veterans EHR System, the INSIGHT Clinical Research Network (CRN) database, the Longitudinal Patient Database for General Practice, the SAIL Databank, the Clinical Practice Research Datalink (CPRD), and NHSDigital, with some allowing linkage to mortality, socioeconomic, registry and audit data.^1–17^

Determining exposures, outcomes and covariates is central to EHR research^18,19^ through generation of medical and drug “codelists” for an overarching clinical definition. Unfortunately, both methodology and reporting vary, forming sources of misclassification bias when ascertaining conditions and prescriptions. Methodology may not involve clinician review, manifesting in exclusion of necessary codes alongside inclusion of inappropriate codes. The Reporting of studies Conducted using Observational Routinely-collected health Data (RECORD) statement calls for EHR studies to provide “complete list[s] of codes and algorithms used to classify exposures, outcomes, confounders, and effect modifiers…considering the risk of misclassification bias…authors should provide sufficient detail to make…research *reproducible*…[and] *risk of bias* apparent”(emphasis added).^20^

Calls regarding transparency and bias extend beyond making codelists freely accessible in repositories. It also requires making methods for their curation freely accessible, systematic and standardizable yet malleable, incorporate clinical input, and designed around proactively considering when bias can manifest upon subsequent application to cohorts. Malleability within codelist design can make reproducibility and generalization to other contexts and databases possible, allowing consistent definition(harmonization) of phenotypes.^13,21^ Malleability facilitates researchers to adapt and reuse others’ codelists for their study and, conversely, facilitates researchers to contribute their codelists to others’ studies appropriately.^22^ Literature on drug codelists has been primarily high-level,^23,24^ underlying steps for its generation of the codelists themselves not usually described.

### Aim

Given unique considerations for prescriptions, we developed a standardizable, reproducible method for creating drug codelists that incorporates end-to-end clinical expertise, considers missing data in search “attribute” variables, considers fluctuating drug status, and is adaptable to other studies and databases. We then operationalize the methodology to generate a codelist for oral drugs for hypertension and heart failure, and a disease-specific codelist, all inhaled therapies for COPD. We also apply the codelists to a sample cohort, according to clinical and study-specific considerations.

## METHODS

### Defining phenotypes, value sets, and ontologies

Common to both medical and drug codelist generation is establishing the single overarching clinical definition (analogous to a “phenotype” for medical codelists), premised by a database’s underlying drug hierarchical ontology (eg, the British National Formulary, BNF, the Anatomic Classification System, ATC, the VANDF (US National Drug File) and RxNorm).^25–28^ Within this definition are “value sets”, a “uniquely identifiable set of valid concept representations” for “possible values of a coded data element in an information model.”^29^ Depending on purpose and study context there may be one or multiple value sets. Ideally, searches to identify all possible codes are conducted purely using the drugs’ chemical terms or the database’s ontology, but missing data can prevent this.

### Our methodology

Our methodology for generating drug codelists has seven steps (summarized in Figure 1) and is available on our GitHub. It is based on our medical codelist methodology, available on our GitHub.

**Figure 1.**
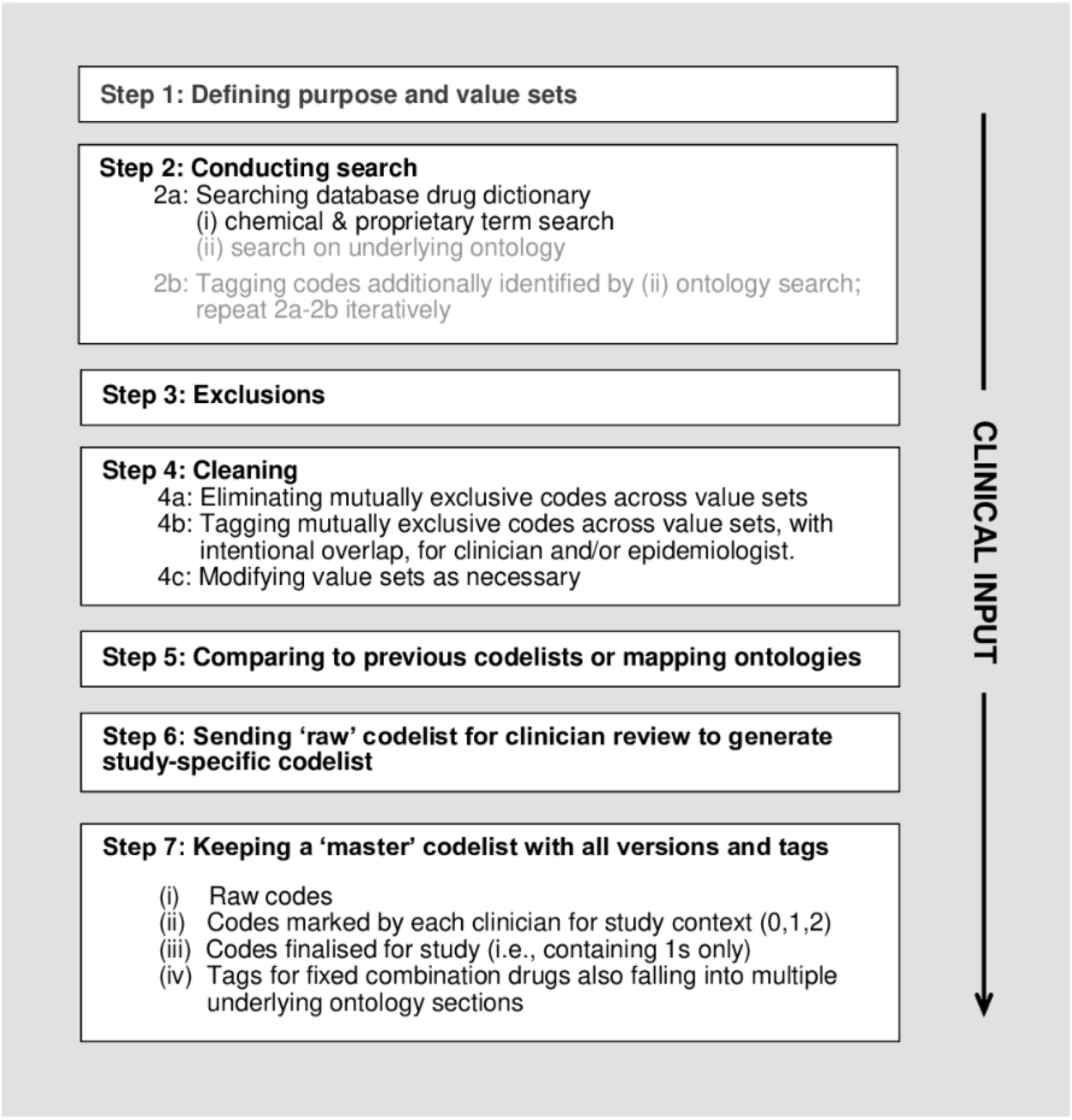
Summary of methodology for generating a product codelist. Note: Steps in grey font may be database-dependent and modified as necessary given missing data.

#### Step 1: defining purpose and value sets

The first stage is to identify value sets consisting of drug class(es) based on an underlying ontology and within classes a list of drugs with corresponding chemical and potentially multiple proprietary names. The route(s) of administration (eg, oral, parenteral) should be specified. The aim may be to produce a broader codelist permitting modification for various contexts, or a study-specific codelist.

We consider clinical input regarding chemistry to improve search efficiency. Searching on common compounds, active or blocking groups, or side chains such as -*nitrate -arginine -hydrochloride - mesilate* is not recommended. Although these suffixes may be listed as part of the drug name, they are not the chemical-of-interest and may lead to inefficiently large search outputs.

Premising step 1 is confirming the database’s ontology to systematically collate a list of all possible terms (ie, the BNF for UK databases) and reliable resource to facilitate reproducibility of collation. A user-friendly BNF resource is the OpenPrescribing^30^ interface utilizing raw data from UK National Health Service Business Services Authority(NHSBSA).

#### Step 2: conducting the search

The second step is to search based on the collated list, using wildcard (*) characters to pick up terms in any location within a string.

##### Step 2a: Search the database drug dictionary

Before the search, we import the database’s drug “dictionary” text file. We import all “attribute” variables searched upon as strings.

We search the browser in 2 stages:

i. For each drug within value sets, search by the chemical and all proprietary names, for a string match on the associated “attribute” variables.
ii. Search on underlying ontology, considering syntax with slashes(/) as medicines may be indicated for multiple conditions and located in chapters not primarily of interest but still used for the indication-of-focus (eg, searching for sildenafil (parenteral route) may fall into section ontologies for both hypertension and erectile dysfunction).

This automated search nests chemical and proprietary terms within each drug list, with corresponding lists nested within broader value sets (Figure 2), in effect sorting output for (i) by value set.

**Figure 2.**
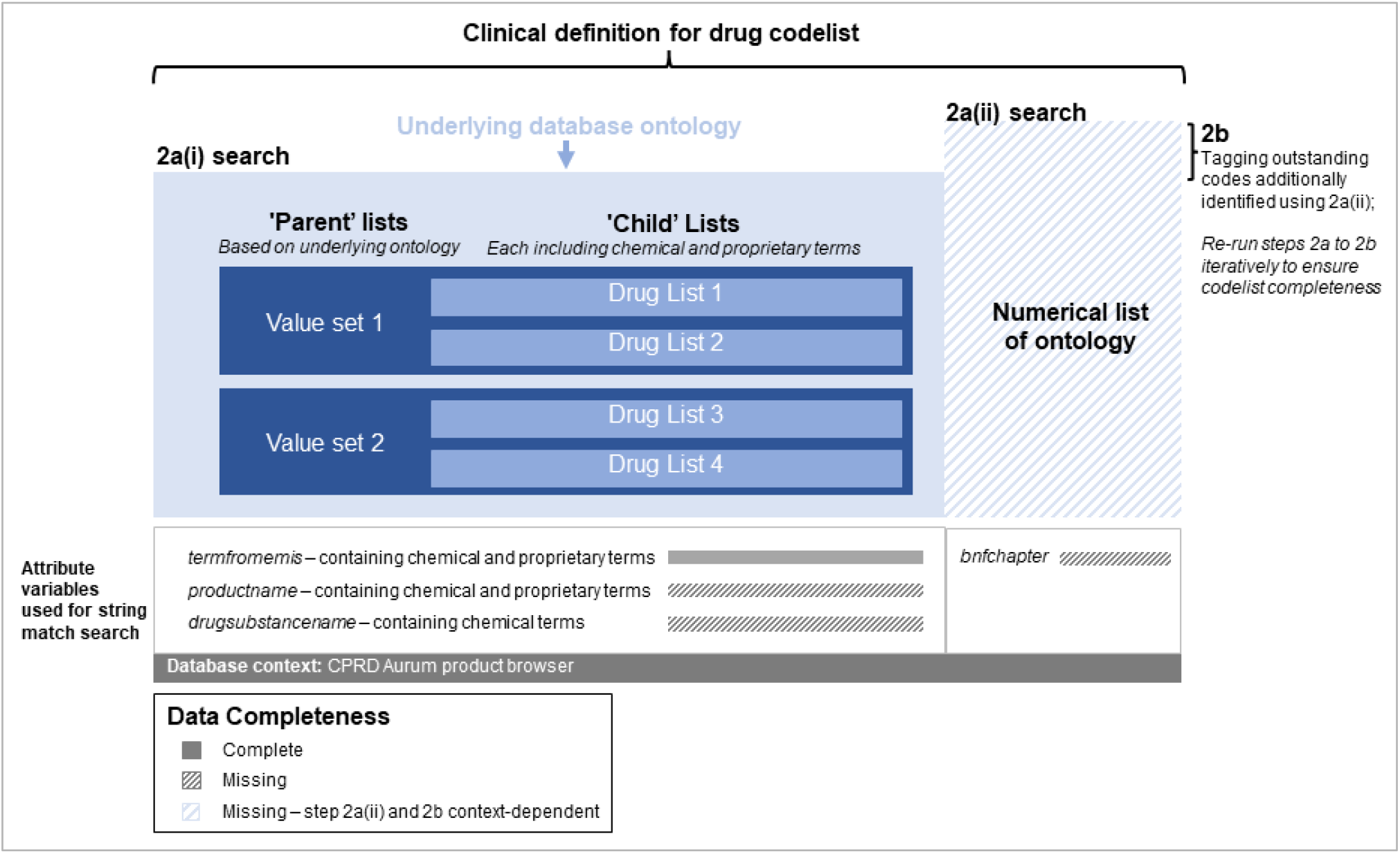
Flow diagram showing the Step 2 search process for drug codes. A main clinical definition for the drug codelist is established, premised by the underlying database ontology. Within each given value set (the “parent” list) are nested “child” lists each corresponding to individual drugs, chemical and proprietary names. The CPRD-specific^5,33^ search attributes are *termfromemis* (i.e., the term from EMIS software) and *productname* (containing chemical and proprietary information) and *drugsubstancename* (chemical information). Due to missing data within the search “attribute” variables we search on all three variables in Step 2a(i), with an additional ontological code search in Step 2a(ii), checking for search term completeness of the former by comparing the (i) and (ii) in Step 2b iteratively. Therefore, Step 2a(ii) and 2b may be database dependent given missing data. In Stata, parent and child lists take the form of local macros; in R a comparable step would be to name a list of vectors, and nesting the lists as necessary. The full Stata and R code including all drug codelist generation steps is located on our GitHub repository.

##### Step 2b: Tag codes additionally identified by searching on (ii) underlying ontology; Repeat 2a-2b iteratively

We tag outstanding codes not found by searching on chemical and proprietary terms alone, allowing researchers to check if they included all possible terms. Outstanding codes are identified and tagged if the row total for columns with “1” denoting searches (i) and (ii) within 2a sum to least two. If outstanding codes are present, one should add the additional names to the value sets, re-running steps 2a to 2b. Ideally, upon multiple iterations of re-running, there should be an absence of tags, indicating inclusion of all appropriate terms. This additional step may seem redundant but is most important to check codelist completeness.

In place of this step we incorporated an expanded search using Systematized Nomenclature of Medicine-Clinical Terms (SNOMED-CT, a commonly-used international reference terminology) concept IDs to check for outstanding synonym codes fitting our value sets, as recommended for medical codelists.^31^ However, in CPRD, although for clinical events a given SNOMED-CT Concept ID will match with multiple SNOMED-CT description IDs, for prescription events a given SNOMED-CT Concept ID does not match with multiple SNOMED-CT Description IDs. This is because the UK SNOMED-CT drug extension is derived from the Dictionary of Medicines and Devices (dm+d),^32^ with the SNOMED-CT Concept ID being identical to the dm+d code, and CPRD only utilizes dm+d codes, preventing this approach.

#### Step 3: exclusions

Step three consists of, after manual review, excluding codes. This is distinguished from later exclusion based on epidemiological and clinical considerations specific to study context. Elimination may be based on information from drug name, route, and/or formulation. The broad search may pick up different medications with the same active chemical but of an inappropriate route, ie, for a different medical indication corresponding to a different organ system.

We do not eliminate by product identifiers; it is a less transparent method. We do not eliminate by ontology chapter either, not only due to missing data, but anecdotal evidence suggests some drugs may have intended medical indication(s) corresponding to multiple chapters, which cannot be assumed and is not reflected in its ontological classification code.

#### Step 4: cleaning

##### Step 4a: Eliminating mutually exclusive codes across value sets

This step places a temporary tag on codes mutually exclusive across value sets, a possibility given the broad search, eg, even within a single BNF class chapter, active ingredients may overlap among the individual sections.

This tag allows researchers to write code automating the re-sorting process to make these sets exclusive.

##### Step 4b. Tagging mutually exclusive codes across value sets, with intentional overlap, for clinician and/or epidemiologist

This step consists of proactively placing permanent tags on codes corresponding to fixed combination drugs with intentional overlap.

We define “intentional overlap” when one code corresponds to a fixed combination drug consisting of two drug classes (ie, mechanisms of action) such that it resides in multiple ontological sections, eg, hydrochlorothiazide/captopril, a single drug including both diuretic and Renin-angiotensin-aldosterone system (RAAS) components.

Clinician input is considered to provide lists of possible suffixes for the tags, eg, “*azide*” for diuretics, or “*pril*” for angiotensin-converting enzyme (ACE) inhibitors and angiotensin receptor blockers (ARBs).

##### Step 4c. Modifying value sets as necessary

This step allows for value set modification, for example, combining sets into a broader value set upon study context or computational considerations (eg, Stata has macro character limits).

#### Step 5. comparing to previous lists or mapping ontologies

This step is for merging together and comparing current and previous codelist versions, and to merge and map codes labelled under different ontologies(eg, ATC-BNF mapping). Comparison facilitates correct categorization and possible identification of outstanding codes. Beyond completeness, mapping allows harmonization and reproducibility to other database contexts.^13,21^

This leads to the “raw” codelist that is not study-specific, and ready for adaptation to a cohort through clinical review.

#### Step 6. sending the “raw” codelist for clinical review to generate the study-specific codelist

Clinician(s) review the “raw” codelist and each code (observation) is labeled as the following

“certainty” categories:

0 = “clear exclusion”

1 = “certainty” to include

2 = “uncertainty” if to include for future sensitivity analyses

Review by one clinician trained in epidemiology and familiar with using the database is necessary, *at least*. When placing certainty categories, clinicians may consider:

− *cohort-of-interest* specific to the organ system. Multiple clinicians are necessary if multiple organ systems are involved, ie, multimorbidity
− operationalization of codes in clinical settings and patient behavior or prescription commonality within context
− *database characteristics* (eg, drug with a substantially low number of issues may have low frequency of prescription events when later applied to the cohort)

An additional step to resolve discordances may be required if multiple clinicians. Step 6 adapts a publication’s clinical review methods for generating medical codelists.^18^

#### Step 7. keeping a “master” codelist

Researchers should keep a “master” codelist with all versions and tags for reasons pertaining to malleability: to allow adaptation for sensitivity analyses and allow generalization to other and harmonization and across study contexts.

It should contain:

i. raw codes – all codes (not study-specific) initially generated by epidemiologist sorted by ontology
ii. clinical input – columns per clinician (≥1 column with 0/1/2s)
iii. study-specific codes – column for tailoring to context (1s “certainty” only)
iv. tags for individual codes for fixed combination drugs also classified within different ontology chapters

### Operationalizing our methodology

The methodology was applied to generate two codelists, 1) drugs for hypertension and heart failure (BNF Chapter 2.5) and 2) all inhaled therapies for COPD (BNF Ch. 3.1.1-.2; 3.1.4; 3.2) in CPRD Aurum^30,33^

We estimated the number of respective prescriptions among a cohort of patients diagnosed with COPD within the CPRD Aurum in England. Many individuals with COPD require inhaled therapies for reduced lung function and have cardiovascular disease.^34^ In this open cohort, patients were included and started follow-up at latest of 1 January 2010 if they 1) were diagnosed with COPD using validated codes,^35^ 2) at least 40 years, 3) had continuous 1-year GP registration, and 4) data was deemed “acceptable” quality. Follow-up ended on earliest of: 31 December 2019, last collection date, death, or transfer out of GP practice.

We compared output of the following searches, for the codelist and upon application to the cohort:

A. Using our full “gold-standard” methodology, searching on chemical and proprietary terms and BNF codes (chemical and proprietary names on *termfromemis (*(i.e., term from EMIS software), *productname* variables, chemical names only for *drugsubstancename* from the nature of this variable, BNF codes for *bnfchapter*)
B. Using our methodology, but searching on chemical names only (within *drugsubstancename*)
C. Using our methodology, but searching on the BNF code only (within CPRD’s *bnfchapter*)

Outcomes were product codes, drug issues, and prescriptions overall and by value set. Analyses were conducted using Stata v17 (Texas, USA). We wrote final scripts in Stata, translating into R v4.2.0. A summary of each codelist’s purpose and operationalization is in Supplementary Table S1. R and Stata scripts, as well as a full term list for the value sets, are located on our GitHub.

## RESULTS

### Generating the raw codelists

We operationalized the methodology in CPRD to generate two codelists, a cardiovascular codelist for hypertension and heart failure medication and a codelist for inhaled COPD therapies.

We designed value sets around codelist purpose (eg, repository or disease-focused). We nested terms corresponding to each drug within “child” lists, then nesting each drug list into “parent” lists(Figure 2). For example, for the indoramin child list (cardiovascular codelist), we searched for chemical and proprietary terms, nesting this list within the BNF chapter 2.5.4 value set). In the COPD inhalers codelist, because intentions were to distinguish disease-specific drugs, we separated value by type even though chemical compositions overlapped, eg, inhaled corticosteroids (ICS) vs. inhaled corticosteroid-long-acting muscarinic antagonists (ICS-LAMA).

In the CPRD drug dictionary, a given unique identifier (*prodcodeid* variable) can include missing data on its following “attributes” including active chemical ingredients (*drugsubstancename*), ontology (*bnfchapter*), and route (*routeofadministration*)(Table S2 describes missing data in the CPRD drug dictionary.) Ideally, searches to identify all possible codes would be conducted purely using chemical terms or the database’s ontology, but missing data prevented this, while the most-complete *termfromemis* variable lists drugs by *either* chemical or proprietary name. Therefore, in in Step 2a(i) we searched on multiple “attribute” variables: *termfromemis, productname* and *drugsubstancename* (Figure 2).

Prior to producing raw codelists, we excluded 26 codes from the cardiovascular codelist, and 206 codes from the COPD inhalers codelist. This was due to cases where composition was correct but route was incorrect given indication (eg, *cutaneous* minodoxil, *ocular* guanethidine monosulfate, salbutamol *nebulizer solutions*), our string match inadvertently picked up codes for a different purpose or distinct chemical compound (eg, Glutenex from searching “*tenex*”; apra-clonidine from “*clonidine*”), or the term was not part of value sets (eg, ICS-salbutamol codes).

We tagged codes for fixed combination drugs also classified within other BNF chapters. For the cardiovascular codelist, this comprised codes for Ch. 2.2 diuretics and 2.6 antianginal drugs, eg, Lisinopril-Hydrochlorothiazide (diuretic and angiotensin-converting enzyme (ACE) inhibitor). For the COPD inhalers codelist this comprised codes for Ch. 3.3, for Salbutamol-sodium cromoglycate.

After respective exclusions and tags, the raw cardiovascular codelist contained 601 codes of both oral and parenteral routes, and for inhaled COPD therapies, 259 codes. For the COPD inhalers codelist, after subsequently merging with a previous codelist mapped to NHSBSA TRUD ATC-BNF ontology mapping files, this led to a final count of 472 codes. Of these codes, 77 were new codes not in the previous codelist; 13 outstanding codes were from the previous codelist not in the new codelist. Most new codes were ICS- or short-acting beta-agonist (SABA)-based (Table S5).

### Clinical review

The first clinician, a respiratory consultant and epidemiologist, removed codes for drugs not part of value sets: for the cardiovascular codelist, nine for Selexipag and one for Sodium Nitroprusside from searching on BNF ontology(Step 2a,ii). There was concern such prescriptions were rare given few issues and it being less likely these infrequently-used, new, or discontinued drugs were prescribed in the COPD cohort. For the COPD inhalers codelist, we removed three codes (0s) consisting of an ambiguous term for route – “liquid” or “solution”– potentially corresponding to nebulized therapies. We retained and tagged one code for pediatrics.

For the cardiovascular codelist, 33 codes of the parenteral route were given 0s as this route is not typically prescribed in UK primary care, leaving oral medications. A second clinician, a cardiologist, reviewed the 1s, agreeing on all codes.

### Final codelists

The final cardiovascular codelist had 568 codes for oral medications (Figure 3;Supplmentary TableS3), including the 66 and 28 product codes tagged for overlap with Chapter 2.2 and 2.6. The value set with greatest count was for drugs targeting RAAS (Ch. 2.5.5, N=375), whereas 2.5.3 and 2.5.8 were the smallest (N=4, N=2 respectively).

**Figure 3.**
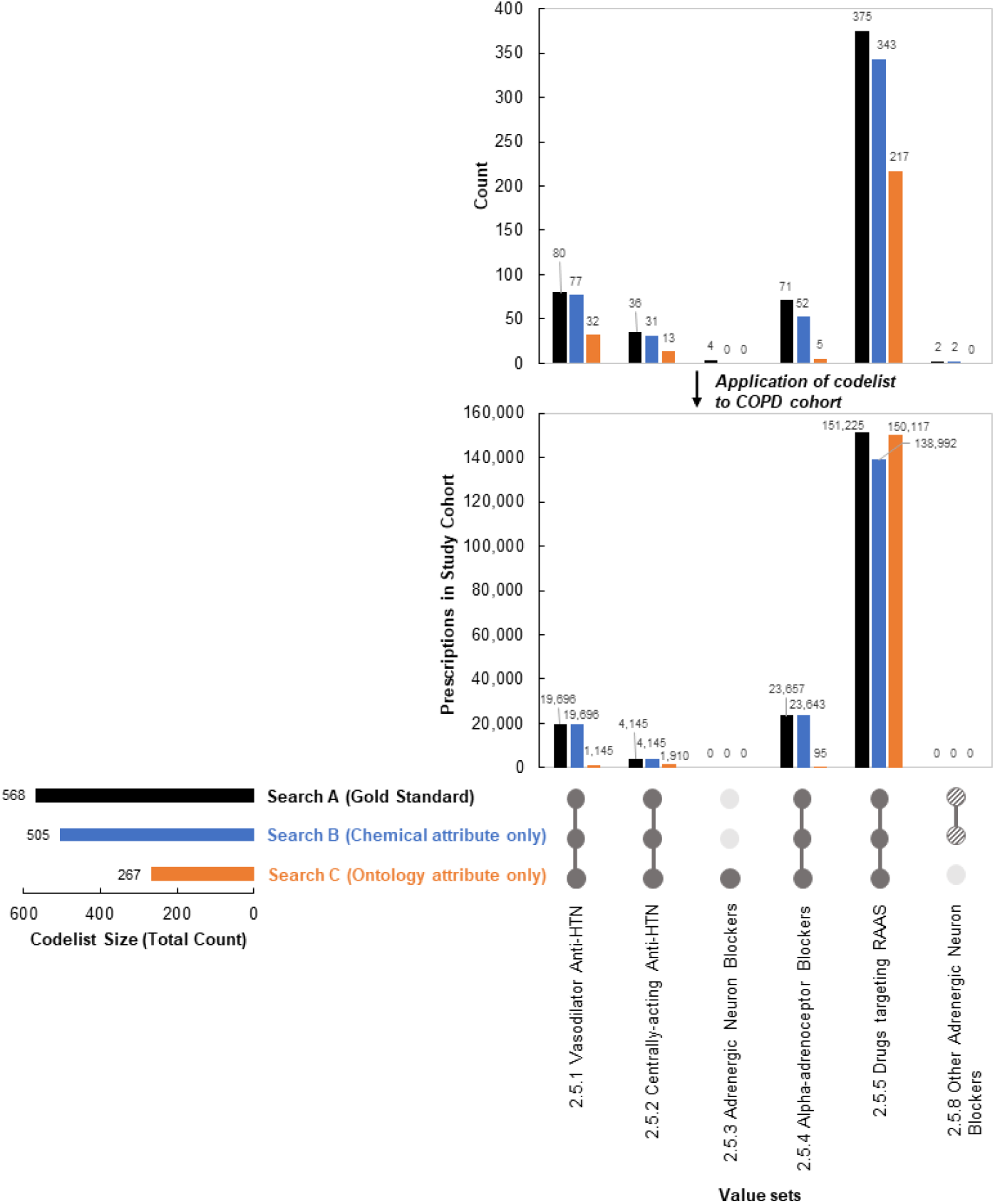
Comparison of codelist output by search type for the cardiovascular codelist. Adapted from an UpSet plot design.^43,44^ RAAS=Renin-angiotensin-aldosterone system. Results refer to post-clinicians’ input. Counts derive from codelist generation, prescribed patients determined after codelist applied to COPD cohort. Prescriptions by value set are mutually exclusive as some patients are prescribed drugs of different classes across value sets, eg, prescribed drug falling into “at least” one subsection. Partially shaded dots for Ch. 2.5.8 refer to presence of codes in codelist, but absence upon application to cohort to determine prescriptions. Search A refers to the use of our methodology, searching *termfromemis, productname, drugsubstancename*, and *bnfchapter* variables. Search B refers to the use of our methodology, but searching on *drugsubstancename* variable, only. Search C refers to the use of our methodology, but searching on *bnfchapter* variable, only (Search C). Refer to Supplementary Table S3 for full data.

The final COPD inhalers codelist had 456 codes(Figure 3;Supplmentary TableS3). The largest value sets were for ICS and ICS-long-acting beta-agonists(LABA) (N=201, N=71, respectively).

### Applying the codelists to find prescriptions

We applied the codelists to a cohort of 335,931 patients diagnosed with COPD according to study population considerations and clinical input.

For the cardiovascular codelist, within the decade follow-up, 165,150 patients (49.2% of cohort) were prescribed at least one of the drugs (Figure 3;TableS3). As in the case with count, the value set for chapter 2.5.5 had the greatest number of patients prescribed (N=151,225, 45.0% of cohort). Chapters 2.5.3 and 2.5.8 did not have prescriptions.

For the COPD inhalers codelist, we determined 317,963 patients (94.7% of cohort) prescribed at least one of the drugs (Figure 4; TableS4). Counts and prescriptions followed different patterns. Whilst ICS had greatest count (N=213 codes), SABA had the most prescriptions (N=297,966; 88.7% of cohort).

**Figure 4.**
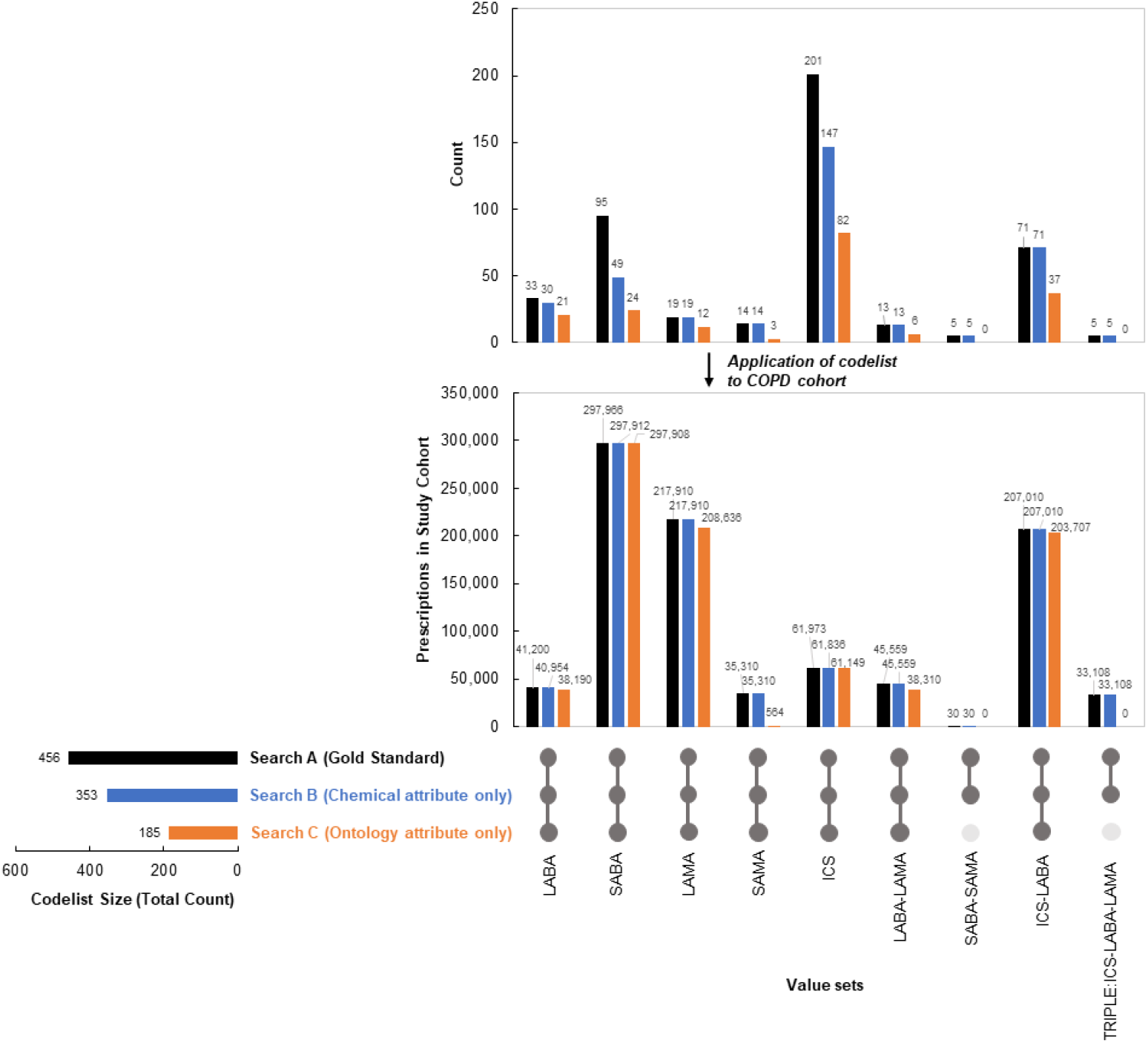
Comparison of codelist output by search type for the COPD inhalers codelist. Adapted from an UpSet plot design.^43,44^ Results refer to post-clinicians’ input. Counts derive from codelist generation, prescribed patients determined after codelist applied to COPD cohort. Prescriptions by value set are mutually exclusive as some patients are prescribed drugs of different classes across value sets, eg, prescribed drug falling into “at least” one subsection. Search A refers to the use of our methodology, searching *termfromemis, productname, drugsubstancename*, and *bnfchapter* variables. Search B refers to the use of our methodology, but searching on *drugsubstancename* variable, only. Search C refers to the use of our methodology, but searching on *bnfchapter* variable, only (Search C). Refer to Supplementary Table S4 for full data.

### Comparing to restricted searches

We compared output of our full “gold-standard” searches (A) to two restricted searches still using our methodology but searching on (B) chemical terms within *drugsubstancename* and (C) BNF ontology within *bnfchapter*.

For the cardiovascular codelist, search B identified 505 codes and 155,678 patients prescribed (46.3% of cohort) at least one of the drugs across value sets. Search C identified 267 codes and 150,669 patients (44.9% of cohort) prescribed at least one of the drugs across value sets (Figure 3; TableS3).

For the COPD inhalers codelist, search B identified 351 codes, and 317,957 patients prescribed (95% of cohort) at least one of the drugs across value sets. Search C identified 185 codes and 315,749 patients (94% of cohort) prescribed at least one of the drugs across value sets (Figure 4;TableS4).

The percent increase in output from searching on BNF ontology only (C) to the gold-standard (A) was the most pronounced (cardiovascular codelist: 113% and 9.6% increase in codes and prescriptions, respectively; COPD codelist 147% and 0.7%). However, we observed marginal increases when comparing Search A to searching solely on chemical information (B) (cardiovascular codelist: 12.5%, 6.08% increase respectively; COPD codelist 29%, 0.0002% increase respectively).

Considering restricted searches by value set, there were absent or marginal increases in prescriptions in some sets upon using Search A (close to 0%), but remarkable increase in others, particularly for C (up to 24802% BNF 2.5.4; up to 6161% for SAMA). Search B led to higher output compared with C, except for Ch. 2.5.5. Here, greater counts in B vs. C did not translate to greater prescriptions (N=343 counts, N=138,992 prescriptions for B; N=217 and N=150,117 for C).

## DISCUSSION

### Summary

We developed a standardizable, reproducible method for creating comprehensive drug codelists using a semi-automated process incorporating end-to-end clinician expertise, considering missing data and fluctuating status, and centered on adaptability to other studies and databases.

We applied the methodology to generate two codelists that were implemented on a sample cohort of patients with COPD in CPRD Aurum, according to study considerations and clinical review.

### Evaluation

Searching only on BNF ontology (C) was insufficient, missing the most prescriptions in CPRD. Given similar results for searches A and B, there may be a marginal opportunity cost between searching on all search variables versus chemical terms alone, depending on drugs desired (ie, restricting to a portion of the value sets, eg, a short-acting muscarinic antagonist (SAMA)-only codelist; or drugs in 2.5.4 only), and cohort size (eg, rare vs. common disease). Value set Ch. 2.5.5 results could be explained by, despite fewer counts, saturation of prescriptions among particular codes, upon codelist application to the sample study cohort.

Although we focus on a given cohort-of-interest with CPRD as our data source, methods center around addition of new information (eg, drugs, proprietary names), completeness, and context-specific adaptations, applicable to other databases and underlying ontologies.

### Methodological recommendations

We recommend the full, “gold-standard” method (A) for comprehensiveness and reaching statistical power (eg, studies where codelist defines cohort or exposure, propensity scoring on complete data). In our database context which contained missing data in the drug dictionary, a search on multiple “attribute” variables was warranted, but this may not be the case for other data sources with data completeness.

If the aim is to produce a broader codelist (ie, higher specificity, lower sensitivity) permitting modification for various contexts, we recommend limiting *a priori* exclusion criteria in step 1; rather, designing sets around underlying taxonomy. If the aim is disease-specific, value set generation should be clinician-led but still designed to permit malleability for different studies, ie, single/fewer classes (eg, statins only). To ensure all possible terms are found we recommend steps 2a-2b iteratively.

Comparisons to previous codelists and/or mapping files can pick up additional codes. In studies incorporating multiple databases,^13,15,21^ we recommend merging with ontology mapping files to enable codelist harmonization, such as ATC-BNF mapping.^36^

### Current literature and tools

Complexities of creating medical codelists are described elsewhere.^18,22,37^ In some databases, recorded prescriptions are treated separately from medical events, in separate tables using different coding schemes, necessitating a separate codelist curation approach.^7,33^ Few studies outline methods for codelist development, with the paucity of literature prominent for *drug* codelists where focus has been high-level, covering challenges, assumptions, and principles in health data research, eg, software and analytical techniques, data preparation, and defining periods for drug covariates^23,24,37^ but not on underlying steps for codelist generation. Of literature on medical codelists, focus was on general guidelines for researchers to enhance reproducibility,^18,24,37^ incorporating clinician review to explore codes’ uncertainty^18^ exploring applications of codelists to sample cohorts^18^ and identifying and comparing disease phenotypes based on restricted versus expanded conceptual definitions.^22,31^

We acknowledge the other software algorithmic tools available to systematically identify patients fitting a broader prescription definition in the record, instead of using underlying drug ontologies^38^ as in the case for countries with healthcare reimbursement processes (ie, countries with mixed-market care, social insurance models, or single-payer national health insurance models that exclude universal insurance of drug prescriptions).^39,40^ These “episode” or “drug groupers” assign drugs, services, and procedures to each patient encounter based on a set of criteria.^38,41^

But drawbacks of groupers point to the current state of drug codelist curation not exhibiting a level of standardizability and reproducibility: evidence indicates grouper methods and criteria lack transparency and are heterogenous^38^ and there is criticism of whether these tools are intended for and/or up to research standard.^38,41^ Furthermore, they may be unavailable or irrelevant in countries with national health service models, in cases of multi-database studies with harmonization of drug definitions^13,21^ and when cost prevents researchers’ use.

### Context-specific considerations

We emphasize consideration of the database and context in all study stages, yet emphasize building modifiability into codelist methodology to allow generalization. Modifications may derive from study *nature*, including the period (eg, retrospective with discontinued drugs in-use during the study), cohort-of-interest (eg, patients with COPD where certain cardiovascular prescriptions are contraindicated), and subsequent statistical analyses. In studies with drug covariates, overlaps in drug class could present collinearity, where exclusion of overlapping codes may be required. Our solution was proactively tagging overlapping codes, drawing upon clinical expertise. Codelist tailoring may relate to *database* factors, such as data type(eg, medications recorded in primary care versus claims data) and taxonomy (eg, BNF or ATC codes).

Due to changes in new, existing, and discontinued drugs, and periods-of-interest for retrospective studies, the same codelist may need updating based on older or newer database versions. Using nested lists allows for maintained organization despite these realities. Future methods may consider adding an extra column to the codelist showing “in use” status, although anecdotal evidence suggests applying codes for discontinued drugs to a newer cohort will not pick up prescriptions.

Results on number of patients prescribed may differ depending on the cohort. Before generating final study-specific codelists, clinical input for exclusion may differ by disease, eg, drugs contraindicated in specific patients such the codelist would be adapted to exclude corresponding value set(s).

Prescriptions may change if the codelist is applied to separate disease cohorts, where prescription commonality by disease varies (eg, cohorts with COPD, diabetes, Chronic Kidney Disease, or multimorbidity).

Results on number of prescriptions may differ depending on how codelists are operationalized, ie, as covariates or exposures, part of inclusion, or accounting factors such as duration and frequency^42^ and for combination, open therapies (eg, for SAMA-SABA beyond fixed therapies determined through codelist generation).

Regarding external overlap to other taxonomic sections, the act of tagging is information proactively sought and built into initial steps, allowing for streamlined adaptability, eg, should covariate collinearity arise. When considering adjustment of confounding covariates, covariate collinearity could distort observed exposure-outcome effects. Resolving collinearity through codelist adaptation should include clinical input.

## CONCLUSION

We designed a semi-automated process to generate drug codelists using standardizable and reproducible methodology. Despite database identity, there are special considerations when generating adaptable drug codelists, including fluctuating status, cohort-specific drug indication and exclusions, database-specific underlying hierarchical ontology, and operationalization relating to inclusion and covariate analysis. Regardless, many EHR researchers are not clinicians; supplemental input is necessary. Premising this is a need to make high quality, rigorous methods accessible to all clinical data researchers in a variety of funding and research contexts.

## Data Availability

Data may be obtained from a third party and are not publicly available. Data are available on request from CPRD. CPRD data provision requires purchase of a license, and this license does not permit the authors to make them publicly available to all.

https://github.com/NHLI-Respiratory-Epi/drug-codelist-creation

https://github.com/NHLI-Respiratory-Epi/SNOMED-CT-codelists

## Author contributions

EG led conceptualization, ethics approval, data curation and formal analysis. PS advised on methodology and analysis, and GM and SH provided analytical support. JQ provided respiratory clinical input when generating the two cohort-specific codelists. NP provided clinical input when generating the cardiovascular cohort-specific codelist. SD advised on visualization. AA translated the STATA software code into R. EG drafted the original manuscript. All authors contributed content, and reviewed and edited the manuscript, with EG and JQ approving the final version.

## Patient consent for publication

Not applicable.

## Acknowledgements

The data used are from CPRD obtained under license from the UK Medicines and Healthcare products Regulatory Agency. The data is provided by patients and collected by the National Health Service as part of their care and support.

## Ethics approval

CPRD has NHS Health Research Authority (HRA) Research Ethics Committee (REC) approval to allow the collection and release of anonymized primary care data for observational research [NHS HRA REC reference number: 05/MRE04/87]. Each year CPRD obtains Section 251 regulatory support through the HRA Confidentiality Advisory Group (CAG), to enable patient identifiers, without accompanying clinical data, to flow from CPRD contributing GP practices in England to NHS Digital, for the purposes of data linkage [CAG reference number: 21/CAG/0008]. The protocol for this research was approved by CPRD’s Research Data Governance (RDG) Process (protocol number: 22_002515) and the approved protocol is available upon request. Linked pseudonymized data was provided for this study by CPRD. Data is linked by NHS Digital, the statutory trusted third party for linking data, using identifiable data held only by NHS Digital. Select general practices consent to this process at a practice level with individual patients having the right to opt-out.

## References

1. Xu Z, Wang F, Adekkanattu P, et al. Subphenotyping depression using machine learning and electronic health records. Learn Health Syst. 2020;4(4):e10241. doi:10.1002/lrh2.10241

2. NCATS National COVID Cohort Collaborative (N3C) Data Enclave. COVID-19 Clinical Data Warehouse Data Dictionary: Based on OMOP Common Data Model Specifications Version 5.3. https://ncats.nih.gov/files/OMOP_CDM_COVID.pdf

3. Justice AC, Gordon KS, Romero J, et al. Polypharmacy-associated risk of hospitalisation among people ageing with and without HIV: an observational study. Lancet Healthy Longev. 2021;2(2):e639–e650. doi:10.1016/S2666-7568(21)00206-3

4. WSIC Data Specification, v11. https://www.registerfordiscover.org.uk/uploads/files/1539001703datadictionary.pdf

5. CPRD Aurum Data Specification, v2.8. Published online August 10, 2022. https://cprd.com/sites/default/files/2022-08/CPRD%20Aurum%20Data%20Specification%20v2.8.pdf

6. VA Family of EHR Cohorts (VACo Family). Accessed March 31, 2023. https://medicine.yale.edu/intmed/vacs/

7. Wood A, Denholm R, Hollings S, et al. Linked electronic health records for research on a nationwide cohort of more than 54 million people in England: data resource. BMJ. 2021;373:n826. doi:10.1136/bmj.n826

8. Data. SAIL Databank. Accessed March 31, 2023. https://saildatabank.com/data/

9. Common Data Model (CDM) Specification, Version 6.0. Published online October 22, 2020. Accessed March 31, 2023. https://pcornet.org/wp-content/uploads/2022/01/PCORnet-Common-Data-Model-v60-2020_10_221.pdf

10. Wolf A, Dedman D, Campbell J, et al. Data resource profile: Clinical Practice Research Datalink (CPRD) Aurum. Int J Epidemiol. 2019;48(48):1740–1740g. doi:10.1093/ije/dyz034

11. Healthcare Epidemiology. Duke Department of Medicine. Accessed April 1, 2023. https://medicine.duke.edu/education-and-training/fellowship-programs/infectious-diseases/training-and-curriculum/research-pathways/healthcare-epidemiology

12. XV Report Health Search. Accessed May 24, 2023. https://report.healthsearch.it/

13. Tran TN, King E, Sarkar R, et al. Oral corticosteroid prescription patterns for asthma in France, Germany, Italy and the UK. Eur Respir J. 2020;55(55):1902363. doi:10.1183/13993003.02363-2019

14. Bezin J, Duong M, Lassalle R, et al. The national healthcare system claims databases in France, SNIIRAM and EGB: Powerful tools for pharmacoepidemiology. Pharmacoepidemiol Drug Saf. 2017;26(26):954–962. doi:10.1002/pds.4233

15. Hsieh CY, Su CC, Shao SC, et al. Taiwan’s National Health Insurance Research Database: past and future. Clin Epidemiol. 2019;11:349–358. doi:10.2147/CLEP.S196293

16. Sohal K, Mason D, Birkinshaw J, et al. Connected Bradford: a Whole System Data Linkage Accelerator. Wellcome Open Res. 2022;7:26. doi:10.12688/wellcomeopenres.17526.2

17. Schull MJ, Azimaee M, Marra M, et al. ICES: Data, Discovery, Better Health. Int J Popul Data Sci. 2020;4(4):1135. doi:10.23889/ijpds.v4i2.1135

18. Watson J, Nicholson BD, Hamilton W, Price S. Identifying clinical features in primary care electronic health record studies: methods for codelist development. BMJ Open. 2017;7(7):e019637. doi:10.1136/bmjopen-2017-019637

19. Sydes MR, Barbachano Y, Bowman L, et al. Realising the full potential of data-enabled trials in the UK: a call for action. BMJ Open. 2021;11(11):e043906. doi:10.1136/bmjopen-2020-043906

20. Benchimol EI, Smeeth L, Guttmann A, et al. The REporting of studies Conducted using Observational Routinely-collected health Data (RECORD) Statement. PLOS Med. 2015;12(12):e1001885. doi:10.1371/journal.pmed.1001885

21. Abbasizanjani H, Torabi F, Bedston S, et al. Harmonising electronic health records for reproducible research: challenges, solutions and recommendations from a UK-wide COVID-19 research collaboration. BMC Med Inform Decis Mak. 2023;23(23):8. doi:10.1186/s12911-022-02093-0

22. Jayatunga W, Stone P, Aldridge RW, Quint JK, George J. Code sets for respiratory symptoms in electronic health records research: a systematic review protocol. BMJ Open. 2019;9(9):e025965. doi:10.1136/bmjopen-2018-025965

23. Pye SR, Sheppard T, Joseph RM, et al. Assumptions made when preparing drug exposure data for analysis have an impact on results: An unreported step in pharmacoepidemiology studies. Pharmacoepidemiol Drug Saf. 2018;27(27):781–788. doi:10.1002/pds.4440

24. Denaxas S, Direk K, Gonzalez-Izquierdo A, et al. Methods for enhancing the reproducibility of biomedical research findings using electronic health records. BioData Min. 2017;10(10):31. doi:10.1186/s13040-017-0151-7

25. VANDF (National Drug File). UMLS Metathesaurus. Accessed February 1, 2023. https://www.nlm.nih.gov/research/umls/sourcereleasedocs/current/VANDF/index.html

26. ATC (Anatomical Therapeutic Chemical Classification System). UMLS Metathesaurus. Accessed February 1, 2023. https://www.nlm.nih.gov/research/umls/sourcereleasedocs/current/ATC/index.html

27. RxNorm Technical Documentation. Published online June 24, 2021. https://www.nlm.nih.gov/research/umls/rxnorm/docs/techdoc.html#s1_0

28. British National Formulary (BNF). Published March 29, 2023. Accessed February 25, 2023. https://bnf.nice.org.uk/

29. SNOMED International. 2.2. Value Set. In: Practical Guide to Reference Sets. International Health Terminology Standards Development Organisation; 2023. Accessed March 18, 2023. https://confluence.ihtsdotools.org/display/DOCRFSPG/2.2.+Value+Set

30. Bennett Institute for Applied Data Science, University of Oxford. All BNF sections. OpenPrescribing. Published 2023. Accessed February 9, 2023. https://openprescribing.net/bnf/

31. Elkheder M, Gonzalez-Izquierdo A, Qummer Ul Arfeen M, et al. Translating and evaluating historic phenotyping algorithms using SNOMED CT. J Am Med Inform Assoc JAMIA. 2023;30(30):222–232. doi:10.1093/jamia/ocac158

32. MacKenna B. Difference between BNF, dm+d and SNOMED CT codes | Bennett Institute for Applied Data Science. Bennett Institute for Applied Data Science, University of Oxford. Published November 23, 2022. Accessed March 31, 2023. https://www.bennett.ox.ac.uk/blog/2022/11/difference-between-bnf-dm-d-and-snomed-ct-codes/

33. Clinical Practice Research Datalink. CPRD Aurum May 2022 (Version 2022.05.001) [Data set]. Published online May 9, 2022. https://doi.org/10.48329/t89s-kf12

34. Morgan AD, Zakeri R, Quint JK. Defining the relationship between COPD and CVD: what are the implications for clinical practice? Ther Adv Respir Dis. 2018;12:175346581775052. doi:10.1177/1753465817750524

35. Quint JK, Müllerova H, DiSantostefano RL, et al. Validation of chronic obstructive pulmonary disease recording in the Clinical Practice Research Datalink (CPRD-GOLD). BMJ Open. 2014;4(4):e005540. doi:10.1136/bmjopen-2014-005540

36. NHS Digital. NHSBSA dm+d. TRUD. Accessed February 1, 2023. https://isd.digital.nhs.uk/trud/users/guest/filters/0/categories/6/items/24/releases

37. Davé S, Petersen I. Creating medical and drug code lists to identify cases in primary care databases. Pharmacoepidemiol Drug Saf. 2009;18(18):704–707. doi:10.1002/pds.1770

38. Peterson C, Grosse SD, Dunn A. A practical guide to episode groupers for cost-of-illness analysis in health services research. SAGE Open Med. 2019;7. doi:10.1177/2050312119840200

39. Tikkanen R, Osborn R, Mossialos E, Djordjevic A, Wharton G. International Profiles of Health Care Systems. Published online December 2020. Accessed April 22, 2023. https://www.commonwealthfund.org/sites/default/files/2020-12/International_Profiles_of_Health_Care_Systems_Dec2020.pdf

40. Brandt J, Shearer B, Morgan SG. Prescription drug coverage in Canada: a review of the economic, policy and political considerations for universal pharmacare. J Pharm Policy Pract. 2018;11(11):28. doi:10.1186/s40545-018-0154-x

41. O’Byrne TJ, Shah N, Wood D, et al. Episode-Based Payment: Evaluating the Impact on Chronic Conditions. Medicare Medicaid Res Rev. 2013;3(3):E1–E26. doi:10.5600/mmrr.003.03.a07

42. Whittaker HR, Wing K, Douglas I, Kiddle SJ, Quint JK. Inhaled Corticosteroid Withdrawal and Change in Lung Function in Primary Care Patients with Chronic Obstructive Pulmonary Disease in England. Ann Am Thorac Soc. 2022;19(19):1834–1841. doi:10.1513/AnnalsATS.202111-1238OC

43. Conway JR, Lex A, Gehlenborg N. UpSetR: an R package for the visualization of intersecting sets and their properties. Hancock J, ed. Bioinformatics. 2017;33(33):2938–2940. doi:10.1093/bioinformatics/btx364

44. Lex A, Gehlenborg N, Strobelt H, Vuillemot R, Pfister H. UpSet: Visualization of Intersecting Sets. IEEE Trans Vis Comput Graph. 2014;20(20):1983–1992. doi:10.1109/TVCG.2014.2346248

