## Supplemental Tables for "Determining prescriptions in electronic healthcare record (EHR) data: methods for development of standardized, reproducible drug codelists"

**Supplementary tables**

Supplementary Table S1. Comparing purpose and methods of the cardiovascular codelist and COPD-specific codelist

|  | **Cardiovascular medication codelist: drugs indicated for hypertension and heart failure** | **COPD medication codelist: inhalers indicated for patients with COPD and prescribed in routine primary care** |
| --- | --- | --- |
| Goal/details of codelist | 1 codelist  Route = initially oral or parenteral  Value set adhere to BNF  Parent BNF organ chapter = 1, **Cardiovascular**  Parent BNF class chapter = 1 (2.5) | 1 codelist  Route = specified inhalers at the start  Value set adhere to BNF, but certain sections removed (3.1.3 theophillines, oral route) during value set creation  Parent BNF organ chapter = 1, **Respiratory**  Parent BNF class chapters = 2 (3.1; 3.2) |
| Information on value sets,***** used to generate search lists | **2.5 Hypertension and heart failure**, into subsections:  2.5.1: Vasodilator antihypertensive drugs 2.5.2: Centrally-acting antihypertensive drugs 2.5.3: Adrenergic neurone blocking drugs 2.5.4: Alpha-adrenoceptor blocking drugs 2.5.5: Renin-angiotensin system drugs 2.5.8: Other adrenergic neurone blocking drugs  *keep all separate | **3.1 Bronchodilators**, into medication type  laba (3.1.1)  saba (3.1.1)  lama (3.1.2)  sama (3.1.2)  sama-saba (3.1.4)  lama-laba (3.1.4)  ics-based single, dual, & triple therapies (3.2) |
| **Internal overlap** - instrinsic to codelist design? | **No**  2.5 **Internal** overlap **not central** to codelist design – when later applied to study (exposure, covariate, etc, inclusion criteria, etc.) - we *may* need to modify codelist (organ-specific codelist, may need to be modified later to be disease-specific)  Value sets guided by BNF section for greater malleability later – 2.5.1, 2.5.2…2.5.8 | **Yes** – codes indicating single, dual, triple therapies are separated through value sets  2.5 Internal overlap ***central* to codelist design** – when later applied to study (exposure, covariate, etc, inclusion criteria, etc.) we care about distinguishing between single/compound therapies, regardless, because of how COPD Rx prescribed (disease-specific-codelist)  Value sets guided by prescribing practices and BNF sections |
| **Tag - External overlap** – flag for future studies when incorporate other codelists | 2.5 **External** overlap  - 2.2 diuretics (e.g., Ramipril-azide)  - 2.6 b,α,Ca blockers | 3.1/3.2 **External** overlap  - 3.3 (e.g., Na chromoglicate) |
| Initial/raw review | Remove all routes – except **keeping oral, parenteral**  Keep all chemicals associated with BNF ontology, because not a disease-specific value set. | Remove all routes – except **keeping inhaled**  Remove chemicals not in disease-specific value set.   - exclude chemicals incorrectly classified into ch. 3 (e.g., prednisolone should be in ch. 6, not 3, so shouldn’t be part of this codelist) - exclude ICS-saba - Remove nebuliser-specific inhalation routes; not typically prescribed in primary care nor part of value set |
| Clinician review | Exclude codes for routes or substances not part of value sets:  Exclude parenteral route because this route is not typically given to COPD patients in primary care. Information about parenteral route may be important in future studies, so the codelist may need to be adapted.  Exclude codes with very low issues | Exclude codes for routes or substances not part of value sets:  Exclude codes that likely indicate nebuliser-specific inhalation route, but were ambiguous in wording, i.e., “liquid.”  Exclude codes with very low issues |
| Future study considerations | e.g., manually determining single vs. dual vs. triple – just because X single-drug codes in codelist are listed as single vs. compound, separate drugs are sometimes combined together to generate the desired compound prescription – separate *prodcodeids* (unique identifiers) | |

*A full description of value sets for each codelist is located on the drug codelists [Github repository](https://github.com/NHLI-Respiratory-Epi/drug-codelist-creation) folder.

Supplementary Table S2. Completeness of Data in the CPRD Aurum Product Browser Dictionary, May 2022 version*

| **Descriptor variable** | **Meaning** | **Completeness**  **N=68,993 observations**  **(N, %)** | |
| --- | --- | --- | --- |
| prodcodeid** | Unique identifier | Complete | |
| termfromemis****** | Term used in EMIS EHR system, may contain either chemical term or proprietary term, *but not both* | Complete | |
| drugissues****** | Total issues within all patients in CPRD, i.e., representative of the general population  (numerical) | Complete | |
| bnfchapter | British National Formulary identifier  (numerical) | 59,839 | 86.7% |
| substancestrength | Dosage strength of drug | 53,664 | 77.8% |
| drugsubstancename | Chemical name, i.e., active ingredient | 53,413 | 77.4% |
| formulation | e.g., solution for injection | 50,284 | 72.9% |
| routeofadministration | e.g., ocular, intravenous, cutaneous | 53,946 | 72.8% |
| dmdid | Unique identifier for DM+D  (numerical) | 12,984 | 18.8% |
| productname | may contain either chemical term or proprietary term, but not both | 12,984 | 18.8% |

*using the February 2022 version of the product dictionary for CPRD Aurum, out of 68,993 total observations in the product browser dictionary.

******denotes variable most complete, i.e., without missing data.

Supplementary Table S3. Number of product codes, drug issues, and patients with COPD prescribed (3A) using all terms (gold-standard), and (3B, 3C) restricted search terms, using the clinician-checked codelist for oral drugs for hypertension and heart failure

| **3A.** |  |  |  |  |  |  |  |  |  |  |  |  |  |
| --- | --- | --- | --- | --- | --- | --- | --- | --- | --- | --- | --- | --- | --- |
|  | **Gold-standard †** | | | | | |  |  |  |  |  |  |  |
|  | **Count** | | **Drug issues** | | **Patients with COPD** | |  |  |  |  |  |  |  |
| **Value Set (BNF Subsection)** | N | | N | | N | |  |  |  |  |  |  |  |
| Total, single issue | - | | - | | 165,150 | |  |  |  |  |  |  |  |
| Total, summation of sets | 568 | | 347,343,632 | | 198,723 | |  |  |  |  |  |  |  |
| **2.5.1 Vasodilator antihypertensives** | 80 | | 15,819,775 | | 19,696 | |  |  |  |  |  |  |  |
| **2.5.2 Centrally-acting antihypertensives** | 36 | | 5,804,240 | | 4,145 | |  |  |  |  |  |  |  |
| **2.5.3 Adrenergic neurone blockers** | 4 | | 9,600 | | 0 | |  |  |  |  |  |  |  |
| **2.5.4 Alpha-adrenoceptor blockers** | 71 | | 31,449,185 | | 23,657 | |  |  |  |  |  |  |  |
| **2.5.5 Drugs targeting RAAS** | 375 | | 294,260,826 | | 151,225 | |  |  |  |  |  |  |  |
| **2.5.8 Other adrenergic neurone blockers** | 2 | | 6 | | 0 | |  |  |  |  |  |  |  |
| **3B.** |  |  |  |  |  |  |  | **3C.** |  |  |  |  |  |
|  | **Chemical terms only ‡** (% increase to Gold) | | | | | |  | **BNF chapter only §** (% increase to Gold) | | | | | |
|  | **Count** | | **Drug issues** | | **Patients with COPD** | |  | **Count** | | **Drug issues** | | **Patients with COPD** | |
| **Value Set (BNF Subsection)** | N | *% inc. to Gold* | N | *% inc. to Gold* | N | *% inc. to Gold* |  | N | *% inc. to Gold* | N | *% inc. to Gold* | N | *% inc. to Gold* |
| Total, single issue | - |  |  |  | 155,678 | *6.08%* |  | - |  |  |  | 150,669 | *9.60%* |
| Total, summation of sets | 505 | *12.5%* | 347,082,016 | *0.08%* | 186, 476 | *6.57%* |  | 267 | *113%* | 294,915,770 | *17.8%* | 153,267 | *29.7%* |
| **2.5.1 Vasodilator antihypertensives** | 77 | *3.9%* | 15,809,535 | *0.06%* | 19,696 | *0%* |  | 32 | *150%* | 1,055,176 | *1399%* | 1,145 | *1620%* |
| **2.5.2 Centrally-acting antihypertensives** | 31 | *16.1%* | 5,761,800 | *0.74%* | 4,145 | *0%* |  | 13 | *177%* | 3,219,000 | *80.3%* | 1,910 | *117%* |
| **2.5.3 Adrenergic neurone blockers** | 0 | *max%* | 0 | *max%* | 0 | *max%* |  | 0 | *max%* | 0 | *max%* | 0 | *-* |
| **2.5.4 Alpha-adrenoceptor blockers** | 52 | *36.5%* | 31,334,678 | *0.4%* | 23,643 | *0.06%* |  | 5 | *1320%* | 98,000 | *31991%* | 95 | *24802%* |
| **2.5.5 Drugs targeting RAAS** | 343 | *9.3%* | 294,175,997 | *0.03%* | 138,992 | *8.8%* |  | 217 | *72.8%* | 290,543,594 | *1.28%* | 150,117 | *0.74%* |
| **2.5.8 Other adrenergic neurone blockers** | 2 | *0%* | 6 | *0%* | 0 | *-* |  | 0 | *max%* | 0 | *max%* | 0 | *-* |

*****Table S3 Corresponds to **Figure 3** in the main text. RAAS=renin-angiotensin-aldosterone system. ‘max%’ refers to change from absence to presence of information. Results refer to post-clinicians' input. Count and drug issues derive from codelist generation, prescribed patients determined after codelist applied to COPD cohort. Prescriptions by value set are mutually exclusive as some patients are prescribed drugs of different classes across value sets, as reflected by the differences in total patients when subsections counts are summated, versus total patients prescribed when only counting a single-issue date, e.g., prescribed drug falling into 'at least' one subsection.

**†** Using our methodology, searching *termfromemis, productname, drugsubstancename*, and *bnfchapter* variables (Search A, gold-standard)

‡ Using our methodology, but searching on *drugsubstancename* variable, only (Search B)

**§** Using our methodology, but searching on *bnfchapter* variable, only (Search C)

Supplementary Table S4. Number of product codes, drug issues, and patients with COPD prescribed (4A) using all terms (gold-standard), and (4B, 4C) restricted search terms, using the clinician-checked codelist for inhaled therapies for COPD*

| **4A.** |  |  |  |  |  |  |  |  |  |  |  |  |  |
| --- | --- | --- | --- | --- | --- | --- | --- | --- | --- | --- | --- | --- | --- |
|  | **Gold-standard †** | | | | | |  |  |  |  |  |  |  |
|  | **Count** | | **Drug issues** | | **Patients with COPD** | |  |  |  |  |  |  |  |
| **Value Set** | N | | N | | N | |  |  |  |  |  |  |  |
| Total, single issue | - | |  | | 317,963 | |  |  |  |  |  |  |  |
| Total, summation of sets | 456 | | 274,518,708 | | 940,066 | |  |  |  |  |  |  |  |
| **LABA** | 33 | | 8,974,130 | | 41,200 | |  |  |  |  |  |  |  |
| **SABA** | 95 | | 129,904,303 | | 297,966 | |  |  |  |  |  |  |  |
| **LAMA** | 19 | | 15,330,100 | | 217,910 | |  |  |  |  |  |  |  |
| **SAMA** | 14 | | 5,848,100 | | 35,310 | |  |  |  |  |  |  |  |
| **ICS** | 201 | | 59,445,136 | | 61,973 | |  |  |  |  |  |  |  |
| **LABA-LAMA** | 13 | | 1,542,720 | | 45,559 | |  |  |  |  |  |  |  |
| **SABA-SAMA** | 5 | | 3,120,000 | | 30 | |  |  |  |  |  |  |  |
| **ICS-LABA** | 71 | | 48,952,319 | | 207,010 | |  |  |  |  |  |  |  |
| **TRIPLE: ICS-LABA-LAMA** | 5 | | 1,401,900 | | 33,108 | |  |  |  |  |  |  |  |
| **4B.** |  |  |  |  |  |  |  | **4C.** |  |  |  |  |  |
|  | **Chemical terms only ‡** (% increase to Gold) | | | | | |  | **BNF chapter only §** (% increase to Gold) | | | | | |
|  | **Count** | | **Drug issues** | | **Patients with COPD** | |  | **Count** | | **Drug issues** | | **Patients with COPD** | |
| **Value Set** | N | *% inc. to Gold* | N | *% inc. to Gold* | N | *% inc. to Gold* |  | N | *% inc. to Gold* | N | *% inc. to Gold* | N | *% inc. to Gold* |
| Total, single issue | - |  |  |  | 317,957 | *0.0002%* |  | - |  |  |  | 315,749 | *0.70%* |
| Total, summation of sets | 353 | *29%* | 264,564,622 | *3.8%* | 939,629 | *0.05%* |  | 185 | *147.03%* | 238,418,300 | *15.14%* | 848,464 | *10.8%* |
| **LABA** | 30 | *10%* | 8,950,830 | *0.3%* | 40,954 | *0.60%* |  | 21 | *57.14%* | 8,109,000 | *10.67%* | 38,190 | *7.88%* |
| **SABA** | 49 | *92%* | 121,362,040 | *7.0%* | 297,912 | *0.02%* |  | 24 | *291.67%* | 117,287,000 | *10.76%* | 297,908 | *0.02%* |
| **LAMA** | 19 | *0%* | 15,330,100 | *0%* | 217,910 | *0%* |  | 12 | *58.33%* | 14,170,000 | *8.19%* | 208,636 | *4.45%* |
| **SAMA** | 14 | *0%* | 5,848,100 | *0%* | 35,310 | *0%* |  | 3 | *366.67%* | 1,510,000 | *287.29%* | 564 | *6161%* |
| **ICS** | 147 | *38%* | 58,056,613 | *2.4%* | 61,836 | *0.22%* |  | 82 | *147.56%* | 49,282,300 | *20.62%* | 61,149 | *1.35%* |
| **LABA-LAMA** | 13 | *0%* | 1,542,720 | *0%* | 45,559 | *0%* |  | 6 | *117%* | 1,290,000 | *19.6%* | 38,310 | *18.9%* |
| **SABA-SAMA** | 5 | *0%* | 3,120,000 | *0%* | 30 | *0%* |  | 0 | *max%* | 0 | *max%* | 0 | *max%* |
| **ICS-LABA** | 71 | *0%* | 48,952,319 | *0%* | 207,010 | *0%* |  | 37 | *91.9%* | 46,770,000 | *4.67%* | 203,707 | *1.62%* |
| **TRIPLE: ICS-LABA-LAMA** | 5 | *0%* | 1,401,900 | *0%* | 33,108 | *0%* |  | 0 | *max%* | 0 | *max%* | 0 | *max%* |

*****Table S4 corresponds to **Figure 4** in the main text. RAAS=renin-angiotensin-aldosterone system. ‘max%’ refers to change from absence to presence of information. Results refer to post-clinicians' input. Count and drug issues derive from codelist generation, prescribed patients determined after codelist applied to COPD cohort. Prescriptions by value set are mutually exclusive as some patients are prescribed drugs of different classes across value sets, as reflected by the differences in total patients when subsections counts are summated, versus total patients prescribed when only counting a single-issue date, e.g., prescribed drug falling into 'at least' one subsection.

**†** Using our methodology, searching *termfromemis, productname, drugsubstancename*, and *bnfchapter* variables (Search A, gold-standard)

‡ Using our methodology, but searching on *drugsubstancename* variable, only (Search B)

**§** Using our methodology, but searching on *bnfchapter* variable, only (Search C)

Supplementary Table S5. Summary of new COPD inhalers codelist compared with previous mapped codelist by value set

| **Codelist** | **Value set** | **N (% difference)** |
| --- | --- | --- |
| 77 outstanding from new codelist | ics | 54 (70.1%) |
|  | Ics-laba | 2 (2.60%) |
|  | lama | 2 (2.60%) |
|  | saba | 18 (23.4%) |
|  | triple | 1 (1.30%) |
| 13 outstanding from previous, ATC-BNF TRUD mapped codelist | laba | 6 (46.2%) |
|  | laba-ics | 3 (23.1%) |
|  | saba | 4 (30.8%) |
